# Post-Exposure Effects of Vaccines on Infectious Diseases

**DOI:** 10.1101/19001396

**Authors:** Tara Gallagher, Marc Lipsitch

## Abstract

Many available vaccines have demonstrated post-exposure effectiveness, but no published systematic reviews have synthesized these findings. We searched the PubMed database for clinical trials and observational human studies concerning the post-exposure vaccination effects, targeting infections with an FDA-licensed vaccine plus dengue, hepatitis E, malaria, and tick borne encephalitis, which have licensed vaccines outside of the U.S. Studies concerning animal models, serologic testing, and pipeline vaccines were excluded. Eligible studies were evaluated by definition of exposure, and their attempt at distinguishing pre- and post-exposure effects was rated on a scale of 1-4. We screened 4518 articles and ultimately identified 14 clinical trials and 31 observational studies for this review, amounting to 45 eligible articles spanning 7 of the 28 vaccine-preventable diseases. For secondary attack rate, this body of evidence found the following medians for post-exposure vaccination effectiveness: hepatitis A: 85% (IQR: 28; 5 sources), hepatitis B: 85% (IQR: 22; 5 sources), measles: 83% (IQR: 21; 8 sources), varicella: 67% (IQR: 48; 9 sources), smallpox: 45% (IQR: 39; 4 sources), and mumps: 38% (IQR: 7; 2 sources). For case fatality proportions resulting from rabies and smallpox, the vaccine efficacies had medians of 100% (IQR: 0; 6 sources) and 63% (IQR: 50; 8 sources) post-exposure. Although mainly used for preventive measures, many available vaccines can modify or preclude disease if administered after exposure. This post-exposure effectiveness could be important to consider during vaccine trials and while developing new vaccines.

## Introduction

Since the advent of variolation in the early second millennium, vaccination has been considered a way to prevent healthy individuals from acquiring disease (1). However, in order to implement informed trials and curb future outbreaks and epidemics, post-exposure effectiveness must be better understood. Significant efforts have recently been devoted to developing therapeutic vaccines for treating chronic conditions such as cancer, diabetes, HIV, and obesity (2), but pre-exposure vaccination remains the focus for communicable disease. One notable exception is the rabies vaccine, which has seen near total efficacy in exposed individuals for the past century. The smallpox vaccine also provided well-documented post-exposure prophylaxis (PEP) until the disease was eradicated 1980, with recommendations dating back to the mid-19th century (71). However, uncertainty surrounding both exposure status and length of incubation make post-exposure properties difficult to estimate for any vaccine, and relatively few studies done so for those that are currently available.

The effectiveness of post-exposure vaccination varies widely depending on disease course, both in terms of individual immune response and population-level spread. While pre-exposure vaccination protects uninfected individuals from infection, post-exposure vaccination serves to modify or prevent clinical disease among those who are already infected. As a result, post-exposure trials must operate within a constrained timeframe: participants must be identified and treated between exposure and symptom onset. Measurable benefits may occur if the vaccine stimulates an immune response faster or larger than that provoked by the natural infection alone. For smallpox, the vaccine has been shown to induce antibody response 4 to 8 days before the variola virus, probably because it bypasses the initial respiratory tract stages of a natural infection (3). These response kinetics explain historical evidence of a post-exposure window and provide a basis for comparing surrogate models to humans in the case of reemergence (4). For rabies, though, this explanation has been unable to fully account for the vaccine’s post-exposure mechanisms. Protection has generally been attributed to neutralizing antibodies, but rabies-exposed patients with HIV have been known to remain well despite poor or undetectable antibody levels after vaccination (5, 6).

On the community level, post-exposure measures could be instrumental in reducing disease burden, especially when mass pre-exposure immunization is not feasible. Mathematical models for tuberculosis have estimated that a post-exposure vaccine would initially be more effective at reducing disease incidence compared to a preventative vaccine, although over time, a pre-exposure vaccine would see increasing impact as more uninfected persons were vaccinated and protected against infection (7). One promising post-exposure candidate, the M72/AS01E vaccine, recently exhibited 54.0% efficacy against disease among a latently-infected population (8). While most diseases do not have such a large pool harboring latent infections – nearly a third of the global population carries tuberculosis – post-exposure vaccination has already curtailed smaller outbreaks of varicella (42, 46), hepatitis A (47, 48), and measles (57, 62). These diseases tend to spread widely during outbreaks, but reactive vaccination studies usually operate under accelerated timeframes and have struggled to distinguish pre- and post-exposure effects. One approach involves considering only the impact of vaccination on symptoms occurring before one incubation period following vaccination, which accounts for some of the uncertainty surrounding exposure status and timing. Variable incubation periods further complicate this method, and make it especially difficult to draw conclusions about vaccination delays. As a result, the most robust information results from studies involving a known exposure during a definite interval prior to vaccination, as is the case for most percutaneous exposures or in settings that practice quarantine.

In order to address post-exposure effectiveness across multiple diseases, this study reviews all infections that currently have an FDA-licensed vaccine, plus dengue, hepatitis E, malaria, and tick borne encephalitis, for which vaccines are available in areas outside the U.S. (Table 1). The evidence could be useful in informing treatment guidelines, but also concerns the design and interpretation of more informative vaccine trials. The body of this review evaluates vaccines administered between exposure and symptom onset, but it also discusses the current state of research surrounding therapeutic vaccines. These are a subset of post-exposure vaccines designed to intervene after the onset of clinical disease, and including a brief summary of them illustrates the broader territory of non-preventive, post-exposure vaccination.

**Table 1:** All infections with an FDA-licensed vaccine, plus dengue, hepatitis E, malaria, and tick borne encephalitis, which have vaccines available in areas outside the U.S. (organized by minimum length of incubation period according to CDC.) The interval listed in the third column is the longest delay with at least 3 studies suggesting an effectiveness of >75%. Diseases with evidence from this study are highlighted in green; studies with no evidence say ‘no data.’

| Eligible infection | Incubation period in days (range) | Interval for successful post-exposure vaccination (maximum) | Current ACIP recommendations for PEP vaccine use |
| --- | --- | --- | --- |
| **Cholera** | .08-5 | No data | None |
| **Pneumococcal disease** | 1-3 | No data | None |
| **Anthrax** | Inhalation: 1-6 | No data | 0, 2, 4 weeks |
|  | Cutaneous: 1-7 | No data |  |
| **Rotavirus** | 2 | No data | None |
| **Diphtheria** | 2-5 (1-10) | No data | Toxoid recommended |
| **Haemophilus influenza type b (Hib)** | 2-7 days | No data | None |
| **Meningococcal meningitis** | 3-4 (2-10) | No data | None |
| **Yellow fever** | 3-6 | No data | None |
| **Dengue** | 4-7 | No data | None |
| **Influenza** | 5-7 | No data | None |
| **Japanese Encephalitis** | 5-15 | No data | None |
| **Typhoid** | 6-30 | No data | None |
| **Pertussis** | 7-10 (4-21) | No data | None |
| **Malaria** | 7-31 | No data | None |
| **Poliomyelitis** | 7-21 | No data | None |
| **Tick-borne encephalitis** | 8 (4-28) | No data | None |
| **Tetanus** | 10 (3-21) | No data | Toxoid recommended |
| **Measles** | 11-12 (7-21) | ≤3 days | ≤3 days |
| **Smallpox** | 12.5 (7-17) | For secondary attack: Insufficient data | n/a (case-by-case decisions) |
|  |  | For fatality: ≤3 days |  |
| **Chickenpox/Varicella** | 14-16 (10-21) | ≤3 days | ≤5 days |
| **Human papillomavirus (HPV)** | 14-240 | No data | None |
| **Mumps** | 16-18 (12-25) | Insufficient data | Vaccine recommended, no timing provided |
| **Rubella** | 17 (12-23) | No data | None |
| **Hepatitis A** | 28 (15-50) | ≤14 days | ≤14 days |
| **Rabies** | 30-90 | ≤5 days (+4 boosters) | 0, 3, 7, 14 days |
| **Hepatitis E** | 42 (14-63) | No data | None |
| **Hepatitis B** | 120 (45-180) | Infants: ≤1 month (+2 boosters) | Infants: routine vaccine series |
|  |  | Adults: Insufficient data | Adults: vaccine series |
| **Tuberculosis** | Weeks to years | No data | None |

## Methods

### Strategy

We searched PubMed for clinical trials or observational studies pertaining to all FDA-licensed and their effectiveness after exposure. Search terms for each disease included but were not limited to:

- [disease] postexposure vaccine
- [disease] postexposure vaccination
- [disease] post-exposure vaccine
- [disease] post-exposure vaccination
- [disease] vaccine after exposure
- [disease] vaccination after exposure

Studies concerned with animal models, serologic testing, or pipeline vaccines were excluded from this analysis. In order to ensure demonstration of a post-rather than pre-exposure effect, we limited consideration to secondary cases either confined to the first incubation period after vaccination or otherwise attributed to exposure before vaccination. Satisfactory evidence of such limitation includes recording the time of vaccination and symptom onset within the vaccinated group, or proving that exposure to the disease ended definitively before vaccination. Most studies compared secondary attack rates (SAR) between vaccinated and unvaccinated groups, but for diseases with high rates of infection after exposure such as rabies and smallpox, studies sometimes looked to fatality proportions among secondary cases as a measure of vaccine effectiveness. Both types of studies were included, along with reactive vaccination trials that met our susceptibility and exposure criteria. In order to determine the exposure ratings (defined in the following section), both authors conducted independent methodological assessments. Any discrepancies were resolved through discussion.

## Definitions

Evidence of post-exposure effectiveness was considered both in terms of secondary attack and fatality outcomes. The effectiveness itself was defined as the relative reduction in outcome risk after having been exposed to a pathogen and subsequently vaccinated against it versus no vaccine or placebo. Because definitions for ‘susceptibility’ and ‘exposure’ vary, the exact descriptions have been compiled for each report in the supplementary documentation. Most studies characterized susceptible individuals as those with negative history of vaccination, but few confirmed with serologic testing. Especially in the case of smallpox, it is thought that prior immunity was likely underestimated (3). An abridged summary of exposure certainty relative to vaccination can be found in Table 2. The analysis groups each study within one of the following four classifications.

**Table 2:**
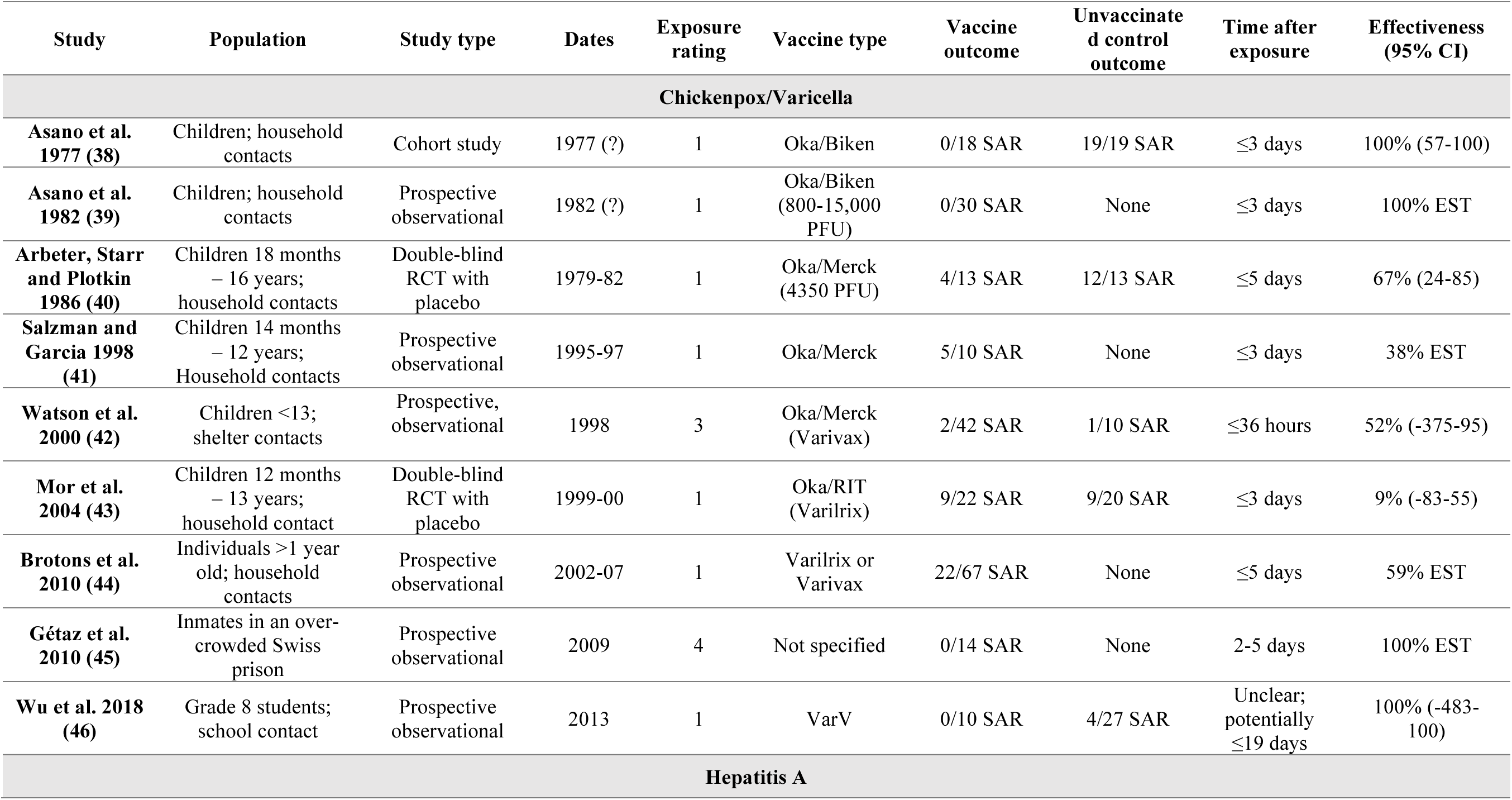
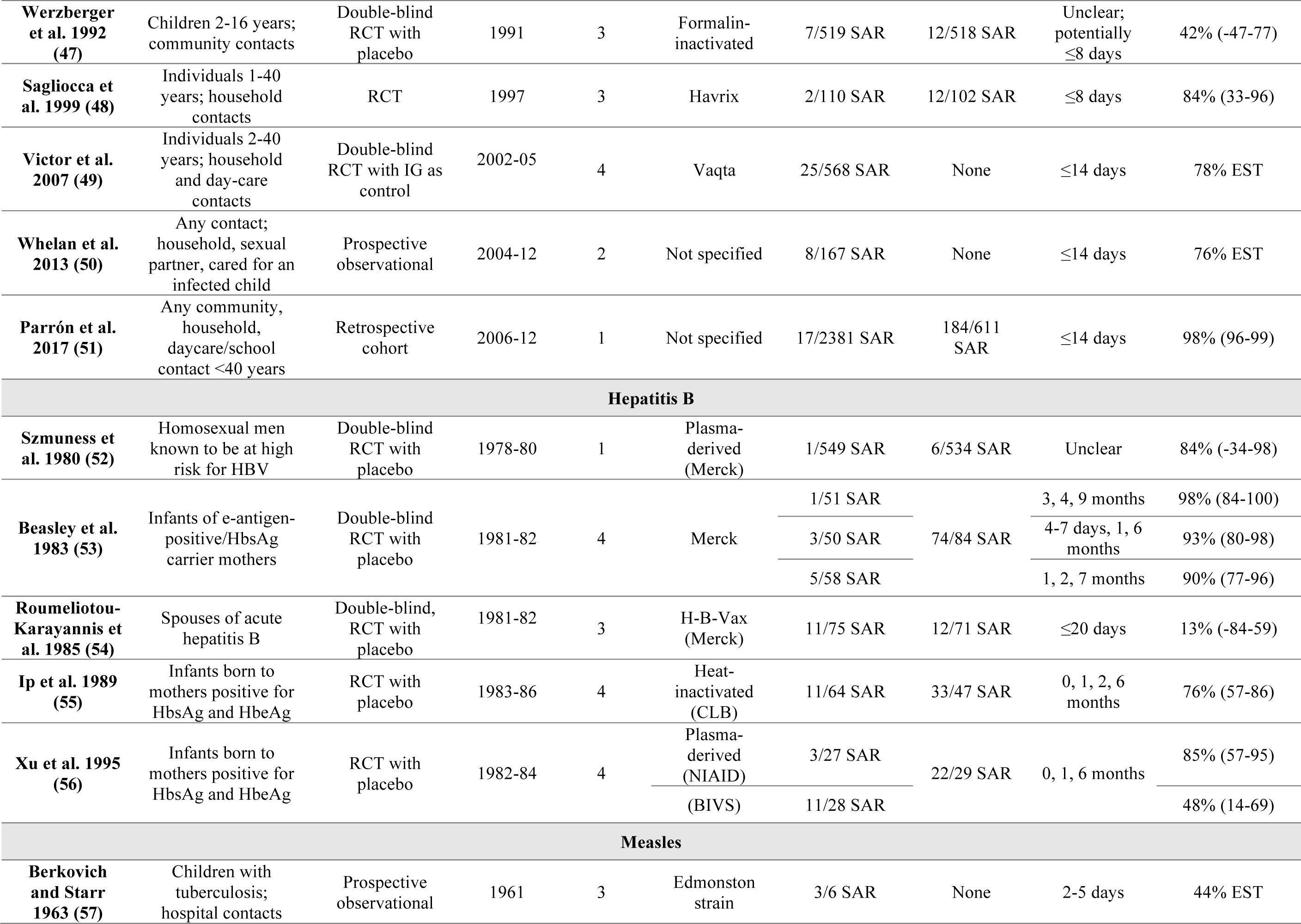
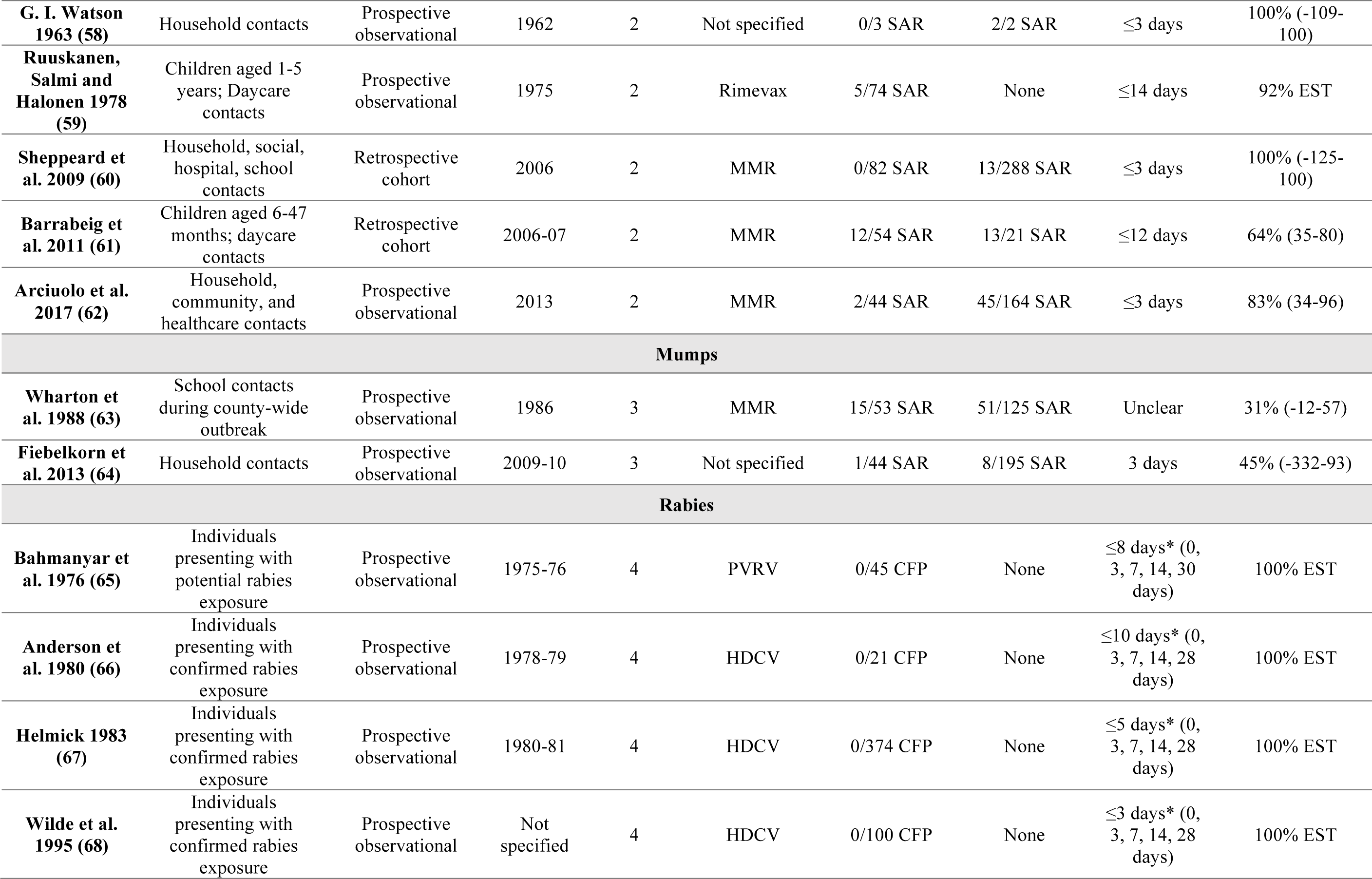
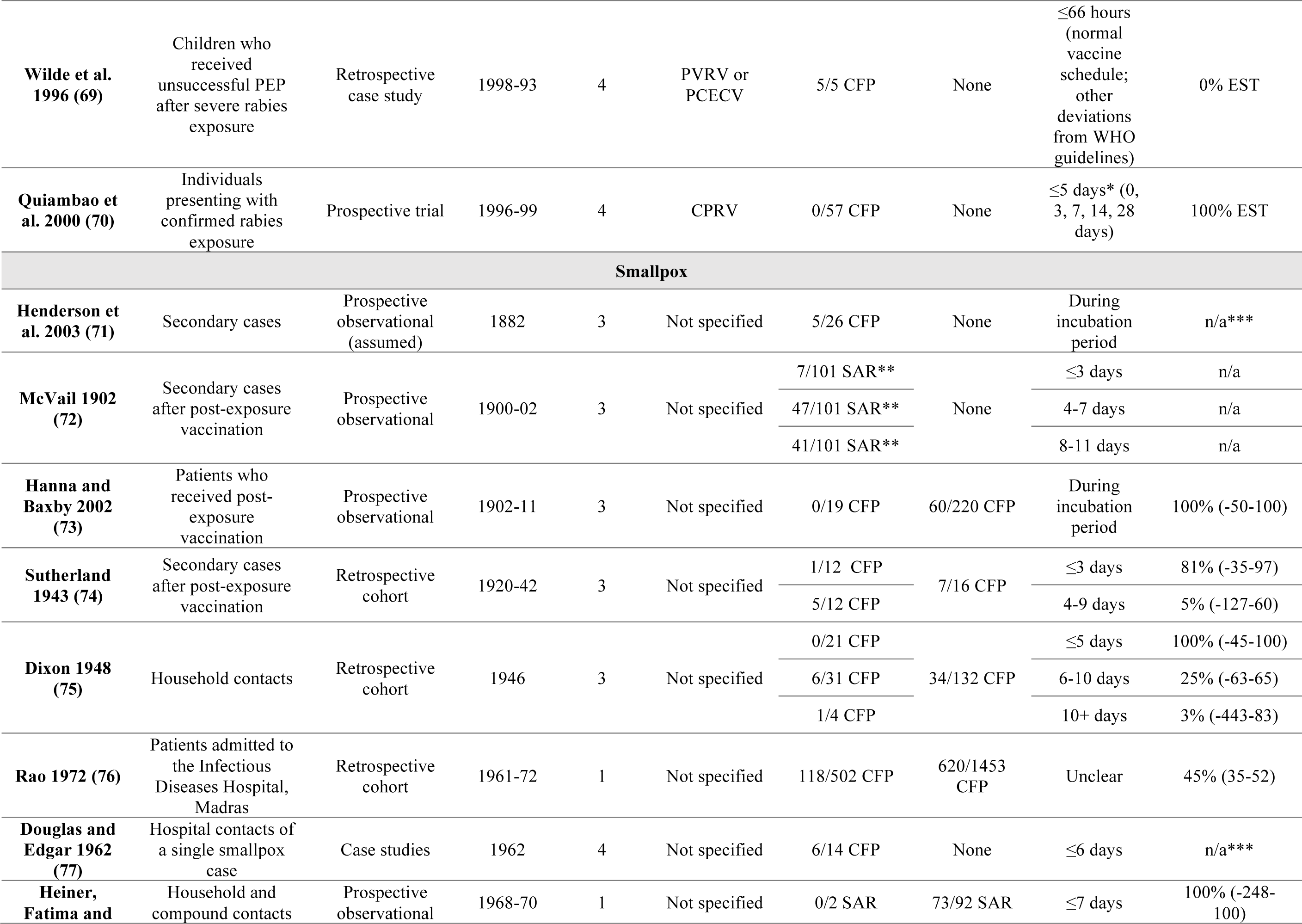
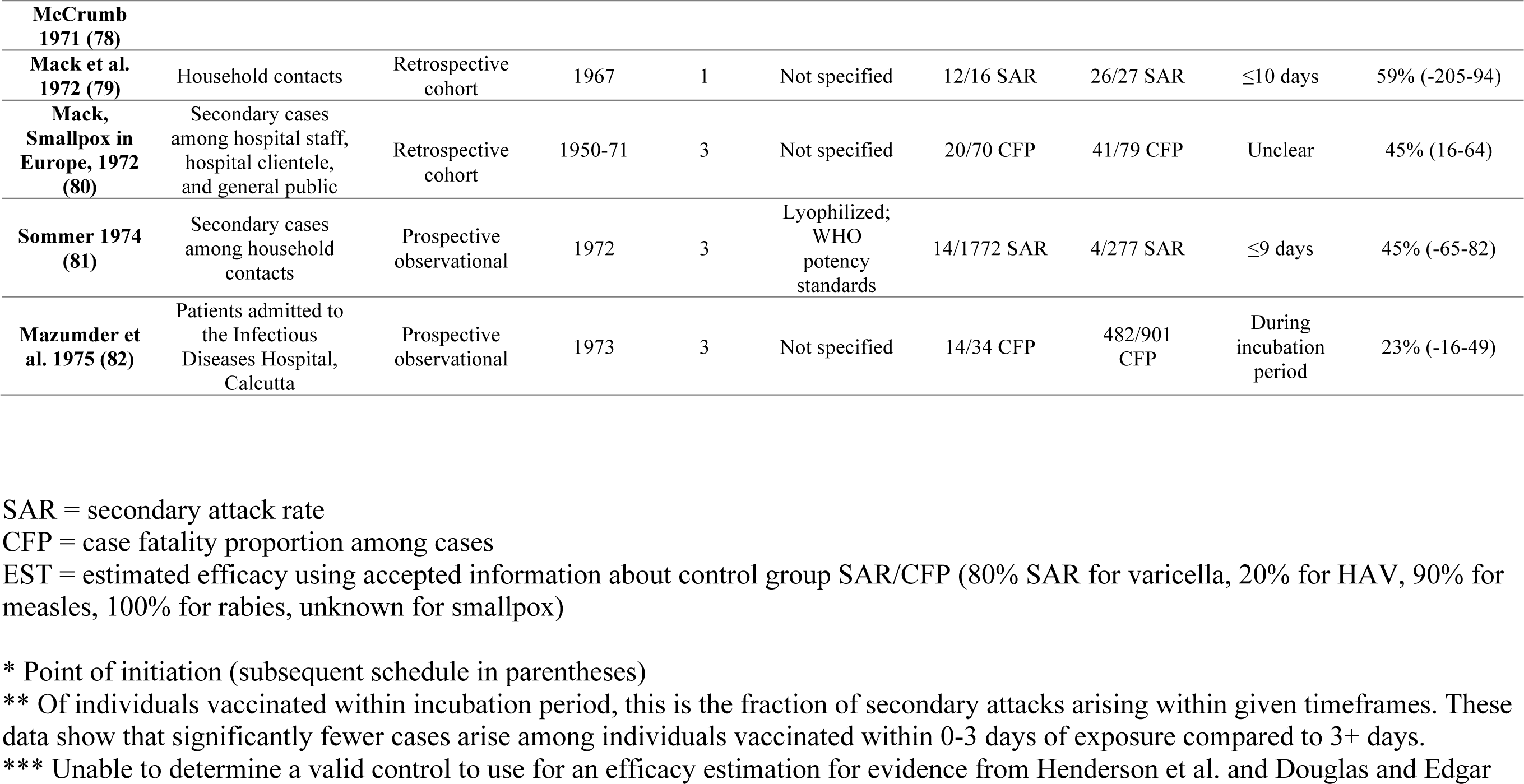
Summary of reports meeting inclusion criteria. Additional notes are listed at the bottom of the table to explain abbreviations. Exposure rating: 1. Exposure is uncertain or ongoing without a close record of timing or timing is followed but without a control group 2. Study indicates a known point of exposure but offers no explanation for their approach 3. Point of exposure can be extrapolated because timing has been followed closely and/or it is likely but not explicitly stated that index patients were isolated 4. There is a clear point where exposure occurs, falling before vaccination

### Exposure rating

1. Exposure is uncertain or ongoing without a precise record of timing or timing is followed but without a control group
2. Study indicates a known point of exposure but offers no explanation for their approach
3. Point of exposure can be inferred with some confidence because timing has been followed closely and/or it is likely but not explicitly stated that index patients were isolated
4. There is a clear point where exposure occurs, falling before vaccination

The vaccines included target infections with an FDA-licensed vaccine, plus dengue, hepatitis E, malaria, and tick borne encephalitis, for which vaccines are available in areas outside of the U.S. These vaccines are referred to as “eligible vaccines” in this report (Table 1). To illustrate the findings from this review, a maximum delay was calculated for each eligible vaccine with evidence, defined as the maximum timespan between exposure and vaccination with at least a 75% effectiveness according to three or more studies. This delay was then compared to the incubation distribution for each disease according to the CDC (Figure 2). For the smallpox vaccine, which has been studied using both secondary attack and fatality proportions as indicators, the included data concern fatality.

**Figure 1:**
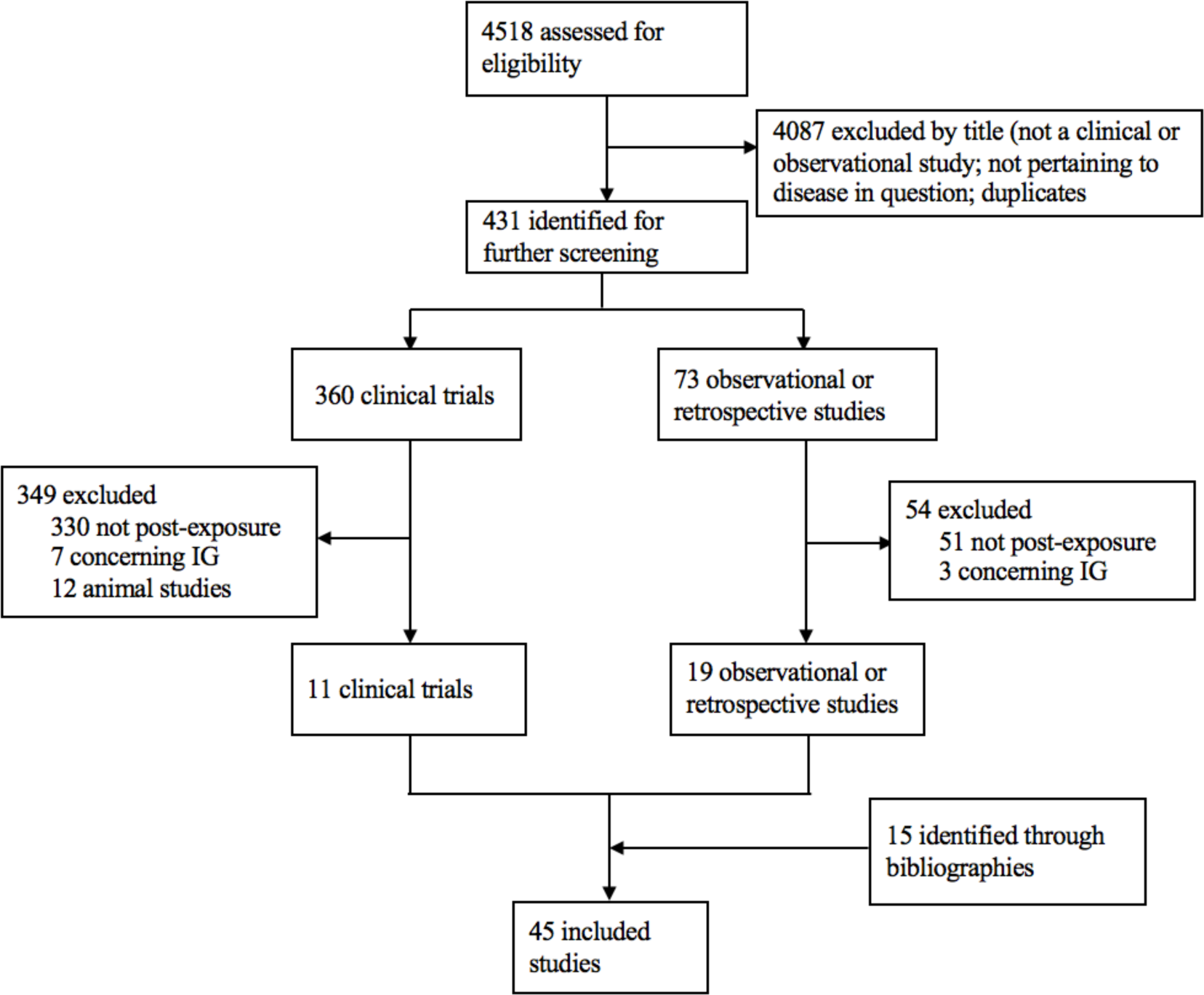
Study selection.

**Figure 2:**
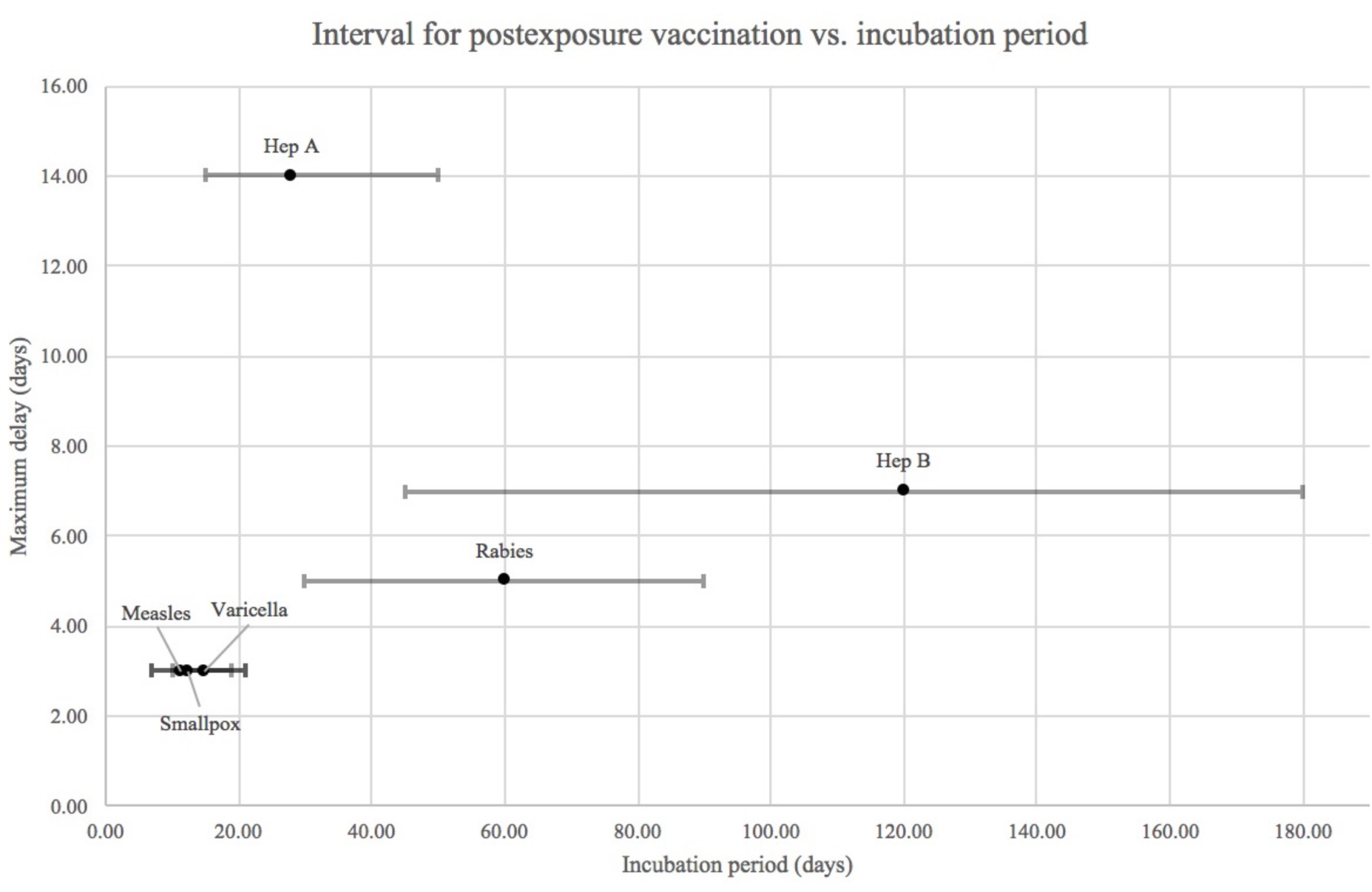
This plot illustrates the post-exposure effect for all eligible vaccines with evidence. The horizontal axis shows the incubation distribution according to the CDC, while the vertical axis shows the maximum delay between exposure and vaccination with at least a 75% effectiveness according to ≥3 studies (at that delay or longer). This effectiveness concerns secondary attack rate for all vaccines except rabies and smallpox, which use fatality as the measured outcome. For example: hepatitis A has an incubation period of 15-50 days with an average of 28, and vaccination is effective anywhere within 0-14 days of exposure according to this review. The two variables have an insignificant correlation (ρ=.76, p=.08).

Where possible, primary data were re-extracted and recorded in Table 2. We elected not to meta-analyze results given that the studies and in particular the inclusion criteria and timing of vaccination were highly heterogeneous for most vaccines, and an “average” effect across such studies would be difficult to interpret. As all point estimates for all vaccines and all studies considered were positive, but not all 95% confidence intervals excluded zero, we simply classified vaccines as those for which statistical evidence of post-exposure protection had been observed in at least one study, and those for which it had not.

## Results

Of the 4518 sources identified, 45 ultimately met inclusion criteria (Figure 1). 434 studies were reviewed by abstract after preliminary exclusion by title, the majority of which were excluded for measuring a preventative vaccine effect (330 of 360 clinical trials; 51 of 73 observational studies). After discarding post-exposure studies focused on immunoglobulin or surrogate models and adding 15 studies found through bibliographies, a total of 14 clinical trials and 31 observational studies reported data for chickenpox/varicella (38-46), hepatitis A (47-51), hepatitis B (52-56), measles (57-62), mumps (63-64), rabies (65-70), and smallpox (71-82). No clinical or observational studies that fit our criteria were located for 21 of the 28 eligible vaccines, although tetanus and diphtheria toxoids have proven to be effective forms of PEP (1).

All but mumps demonstrated statistical evidence of a positive post-exposure effect in at least one study. While the effects measured for smallpox were variable and the median lower than for most other vaccines, reports that stratified by delay after exposure showed a clear relation between prompt immunization and modified disease course. Vaccine effectiveness against secondary attack varied in strength across diseases, with medians of 85% (IQR: 28) for hepatitis A, 85% (IQR: 22) for hepatitis B, 83% (IQR: 21) for measles, 67% (IQR: 48) for varicella, 45% (IQR: 39) for smallpox, and 38% (IQR: 7) for mumps. These medians exclude the efficacies estimated using a historical control (labeled “EST” in Table 1), incorporating only studies with an internal control population. For studies that stratify by vaccination delay (such as the Sutherland (74) and Dixon (75) smallpox studies), the included estimate refers to the shortest post-exposure interval. 15 of the 46 studies considered vaccine effectiveness against fatality – 7 for rabies and 8 for smallpox – and determined median vaccine efficacies of 100% (IQR: 0) and 63% (IQR: 50) for the two diseases respectively.

Within these studies, vaccination timing varies widely and must be a factor when considering these results. According to data in this review, there is an insignificant correlation between the length of incubation and the length of delay between exposure and vaccination that allows for ≥75% effectiveness (ρ=.76, p=.08), but it appears that a longer incubation period allows for a longer delay (Figure 2). Sample populations may also affect vaccine effectiveness: for instance, evidence for varicella and measles tends to focus on children (7 of 9 and 4 of 6 studies respectively), and hepatitis A on individuals under age 40 (3 of 5 studies). Most data for these diseases as well as mumps and smallpox derive from reactive vaccination campaigns, which often saw large enough sample sizes to study. Still, their favorable characteristics sometimes came at the cost of exposure certainty, such as studies conducted in healthcare (44, 62, 76) and school (46, 51) settings that struggled to determine when exposure occurred. Already exposure is difficult to trace and isolate in outbreak settings, and many reactive attempts focused on disrupting an outbreak rather than investigating vaccination as PEP to begin with. Conversely, percutaneous or mucosal exposure to viruses like hepatitis B and rabies is often much simpler to identify and link to infection.

## Discussion

Of the eligible vaccines with relevant post-exposure evidence, all but mumps show compelling evidence of some form of post-exposure protection. Previous reviews have already investigated hepatitis A and B, smallpox, and varicella individually, and while this study is the first to our knowledge to incorporate multiple diseases, its findings align with those targeted reports (9, 10, 11, 12, 13). Using historical smallpox data, Keckler et al. (9) and Henderson et al. (71) conclude that a 3-4 day interval suffices for significant post-exposure protection, while a vaccine administered any time before symptom onset could be advantageous. Expert opinions culled using the Delphi technique corroborated this, estimating that vaccination is 93%, 90%, and 80% effective at preventing smallpox within 0-6 hours, 6-24 hours, and 1-3 days of exposure respectively, and 80%, 80%, and 75% effective at modifying disease among those who develop illness (3). A Cochrane Review of varicella vaccines also determined a 3 day window, but could not locate enough evidence to draw conclusions about vaccine effectiveness beyond 3 days (13). Another review compared perinatal hepatitis B vaccination to placebo or no intervention found a relative risk of 0.28 (.20 to 0.40), even with varying immunization schedules (11).

The findings here agree with those previous reviews and indicate that even common vaccines have properties that have not been fully explored. Of course, post-exposure trials are especially complicated because they involve a pool of infected individuals. The few studies that do exist operate in two main ways, largely depending on how easy it is to identify such a sample. The first approach is to record the time elapsed between vaccination and disease, then infer exposure timing and the window for effective vaccination based on an incubation period estimate. This approach allows for less precise knowledge about exposure timing, and thus lends itself to outbreak situations for diseases like hepatitis A, measles, mumps, varicella, and smallpox. However, each study reconciles these uncertainties differently, so their designs must be considered before comparing their results. For instance, reported events can vary in definition: smallpox onset could refer to either fever (82) or rash (74, 78), which typically develops 2-3 days after the fever.

The second approach is more straightforward and involves restricting consideration to exposure occurring in a distinct period of time before vaccine receipt. Examples include outbreak settings that enforce quarantine (45, 77), or diseases like hepatitis B and rabies where transmission can be linked to specific events. Rabies is unique in that vaccines are typically administered after exposure; however, because current guidelines are known to be successful, deviations larger than the typical 5-day delay have not been studied rigorously (14, 15). Most documented PEP failures tend not to result from vaccine schedule changes, rather from insufficient wound infiltration or rabies immunoglobulin dosage (69). Tetanus also spreads by way of cuts and punctures, although its established treatment protocol has been investigated even less systematically than rabies. The success of post-exposure vaccination by tetanus toxoid has ruled out the option of randomized trials, and the success of preventive vaccination has reduced incidence of tetanus dramatically since the mid-1930s, eliminating most opportunity for case studies (1).

For both approaches, experimental designs create potential sources of variation across studies. Studies isolated exposure with varying levels of certainty, and although no significant pattern emerged between exposure rating and vaccine effectiveness, this rating scale indicates which of them are most likely capturing the desired effects. Other variables include vaccine types and doses; sample sizes, which range from 10 or fewer individuals (41, 57, 58, 69) to over 2,000 (51, 81); prior immunity among participants, which all but a few studies monitored (51, 72, 77); and settings, whether in schools, hospitals, prisons, or households. More detailed information specific to each study can be found in the supplementary documentation.

In addition to these discrepancies, most post-exposure studies assume that post-exposure vaccination itself does not cause symptoms that could be mistaken for mild versions of the illness. If erroneous, this could could lead to an underestimation of vaccine effectiveness by mistaking vaccine-associated rash (for example) for breakthrough infection. One study in healthy (presumably unexposed) children found that 5.9% of MMRV recipients and 1.9% of MMRII/VARIVAX recipients experienced a very mild rash following vaccination, although both groups demonstrated >90% response rates to the vaccine (16); therefore, studies on post-exposure vaccination for varicella and measles may conflate minor adverse events with secondary attacks and underestimate how often the vaccine works. For ethical and logistical reasons, some reports also lack information about important controls: individuals exposed to and untreated for rabies, for example, or individuals who receive a post-exposure smallpox vaccination and are protected from disease. This is the case for all but three historical smallpox reports (78, 79, 81) while the rest include information only about people presenting with smallpox, such as the severity and outcome of their disease.

As for the vaccines without evidence of a post-exposure effect, a likely explanation is that they do not have time to induce an adequate response before clinical disease manifests. The shortest incubation period among diseases with post-exposure protection is 11-12 days for measles, while all but four of the 21 diseases without evidence have incubation periods of 10 days or below (Table 1). For diseases that benefit from post-exposure vaccination, a longer incubation period may permit a longer window for post-exposure vaccination to be effective (Figure 2). Similar biological considerations already inform treatment guidelines when no trials are available: for instance, tetanus toxoid is recommended promptly after exposure because too long a delay would allow additional tetanus neurotoxin to bind to neurons in the peripheral and central nervous system (17). However, post-exposure vaccination against tetanus produces an adequate amount of antitoxin in just 4 to 7 days, leaving a small window for a post-exposure vaccine to outpace the natural, 10-day incubation (1). As discussed previously, the smallpox vaccine has a similar advantage over the natural infection and begins its course a few days ahead, spreading directly to regional lymph nodes and lymphoid organs (3). These results indicate that the success of post-exposure prophylaxis depends upon the timescales of the vaccine as it relates to disease mechanisms. It is important to note, though, that a post-exposure vaccine could still supplement other treatments even if it cannot prevent or modify disease alone. Anthrax infections are one example, and require antibiotics due to a short incubation period and rapid onset. However, because anthrax spores have been known to survive antibiotic prophylaxis, a vaccine should also be administered to counter long-lasting threats (1).

An exception to this generalization is the bacille Calmette-Guérin (BCG) vaccine, which is the only licensed vaccine for tuberculosis and offers no known benefit to individuals with latent infection, despite the fact that latent (asymptomatic) infection can last for years or decades (18). Better defense against tuberculosis will ultimately require a new vaccine, and ideally one that functions both before and after infection. Over a dozen candidates for both priming and boosting have entered clinical trial to date, six of which are designed for individuals with latent or active infections – termed post-exposure or therapeutic candidates, respectively (18). The M72/AS01E vaccine, one of the most promising post-exposure vaccines, recently exhibited 54.0% efficacy among a pool of healthy, *M. tuberculosis*-infected individuals during a two-year follow-up (8). This finding, along with a recent BCG-revaccination trial that saw 45% efficacy within uninfected population (19), has encouraged efforts toward new vaccine strategies for tuberculosis. Future solutions will have to reimagine how vaccines are implemented along the disease course, whether that involves a new way of using BCG or a new vaccine altogether.

In addition to these investigative tuberculosis vaccines, several others in the pipeline may have unexplored post-exposure effects. Filoviruses incubate for a week or longer and thus might be promising candidates are a likely candidate according to the findings of this review. In case reports, the rVSV-ZEBOV vaccine has appeared to alter the course of Ebolavirus (EBOV) following several needlestick exposures in humans (20, 21), and controlled studies have found a 50% post-exposure efficacy in nonhuman primates after receiving a lethal challenge (5). Since disease course is both faster and uniformly lethal in this nonhuman primate model than in humans, the protective effectiveness of the EBOV vaccine could plausibly surpass 50% in humans (22). Preliminary research for Marburgvirus (MARV) indicated that the rVSV-MARV vaccine had similar properties, demonstrating a significant effect if administered within one day of exposure in rhesus macaques (23). In light of the 2013-2016 EBOV epidemic, filovirus countermeasures should remain high priority and systematic plans for gathering evidence should be set in place for the next outbreak.

An intermediate case not considered explicitly in this review is vaccination against herpes zoster disease. This disease, also called shingles, results from the reactivation of latent varicella-zoster virus infection primary, typically symptomatic, varicella disease (chickenpox). Two vaccines are licensed in the US and have been proven effective in preventing herpes zoster (24, 25). While technically a “post-exposure” effect, we have not included this in the main review because it is not an effect in preventing primary symptomatic disease, but rather reactivation.

Classic vaccine efficacy trials are designed to study the preventive effectiveness of vaccination against infection and disease. In most cases, if there were a post-exposure effect of vaccination, it would have little effect on the outcome of such trials, as a small proportion of trial participants would be infected at the time of vaccination. Two classes of exceptions are worthy of note. First is vaccines against bacteria which colonize the respiratory or digestive tract asymptomatically and for which disease is a comparatively rare complication of colonization. For vaccines against such bacteria, the limited evidence available is that vaccination does not terminate the carriage state, but rather reduces the acquisition of carriage and also the probability of disease given carriage (26). If this is the case, then the post-exposure effect should be modest for these vaccines and not complicate estimates of effectiveness. On the other hand, a setting where post-exposure effectiveness could have greater consequences for the interpretation of vaccine trials is in ring-vaccination trials of vaccines against acute viral diseases, such as the Ebola ça suffit! trial that evaluated the rVSV-ZEBOV vaccine against Ebola virus disease in Guinea (27). In such a trial, by design vaccination occurs close to the time of likely exposure to infection, and cases occurring within one incubation period of vaccination (plus one week for vaccine immunity to ramp up) were excluded from analysis as unpreventable by vaccination. On one hand, it is possible that post-exposure effects, such as those observed in nonhuman primates with this vaccine, mean that some cases before that window could be preventable by vaccination, and it would be interesting to analyze the data from before the window in the main analysis to see if there is evidence of such an effect. On the other hand, because incubation periods are variable, it is possible that a window designed to exclude any individuals infected before vaccination would do so imperfectly, leading to some inclusion of a post-exposure effect in the main effect estimate. If this design is used again, it would be valuable to quantify the likely impact of post-exposure effectiveness on estimates obtained from the trial.

With the exception of tuberculosis, most post-exposure benefits discussed so far arise as side-effects of successful preventative vaccines. However, active therapeutic immunization has recently become a focus for chronic, mainly non-communicable diseases. Many of these experimental vaccines induce antibody production, but with the goal of altering a disease that has already begun. Some treat drug abuse by binding to addictive substances like nicotine and cocaine (28), and others block tumor necrosis factor-α (TNF-α), an inflammatory cytokine linked to Crohn’s disease, rheumatoid arthritis, and psoriasis (29). Several active therapy candidates for Alzheimer’s have entered human clinical trials since 2000, mostly to target amyloid β plaques that are thought to be causative agents (30). However, despite promising preclinical results, no significant cognitive benefit has been observed to date (31). A similar narrative characterizes the efforts toward therapeutic HIV-1 vaccines: early optimism because of a slow and relatively well-understood disease progression, followed by decades of research and few positive clinical outcomes (32).

Other therapeutic vaccine candidates induce T cells rather than antibodies, an approach well-suited to treating cancer. In theory, the paradigm mirrors that of classical viral vaccines in that tumor-associated antigens are used to activate T cells, which differentiate and proliferate in order to target the unwanted tumor cells (1). However, despite several promising phase III trials (33, 34), an objective review of cancer vaccine trials found a response rate of less than 4% (35). The U.S. Food and Drug Administration (FDA) has only approved one cancer vaccine, Sipuleucel-T, which targets metastatic prostate cancer and prolongs median survival by 4.1 months (36). Still, the same phase III trial that led to its licensure presented no significant effect on other important disease factors: time to disease progression, tumor regression, long-term immunity. Interestingly, a different licensed vaccine may have the ability to eradicate other malignant tumors, specifically squamous cell carcinomas. In a recent case report, the 9-valent HPV vaccine (Gardisil-9) resolved all of a woman’s tumors within one year of her first injection (37). Although the HPV vaccine has already been linked to preventing cervical cancer and others, its therapeutic properties remain unclear (1).

Therapeutic vaccination represents a promising frontier for disease treatment, but also has implications for how we consider vaccines as a tool for prevention. As evidenced by this report, post-exposure effectiveness has not been fully explored even for common vaccines, and there are several scenarios in which they are important: responding to unpredictable health emergencies, designing new treatments, and interpreting vaccine trials. Especially in outbreak settings, attributing all outcomes to preventative effects could lead to an overestimation of a vaccine’s preventive effectiveness, and under-appreciation of its post-exposure effectiveness. Therefore, post-exposure effects not only concern innovative treatments for exposed or infected individuals, but could also improve how we anticipate and understand the impact of any vaccine. This review represents the current body of evidence for vaccines that are already available, and indicates that there is still much to learn about post-exposure vaccination.

## Data Availability

The data that support the findings of this study are available in the public domain at NCBI - PubMed. Their references are listed within the review.

## Acknowledgment

### Grants and financial support

The project was supported by Grant Number U54GM088558 from the National Institute of General Medical Sciences. The content is solely the responsibility of the authors and does not necessarily represent the official views of the National Institute of General Medical Sciences or the National Institutes of Health.

### Thanks

We thank Dr. Mark Slifka for helpful discussions before this project began.

### Conflicts of interest

ML discloses consulting or honoraria from Merck, Pfizer, Antigen Discovery and Affinivax, and grant funding through his institution from Pfizer. TEG declares no conflict of interest.

